# Corticospinal Responses Following Gait-Specific Training in Stroke Survivors: A Systematic Review

**DOI:** 10.1101/2022.10.28.22281102

**Authors:** Yosra Cherni, Alexia Tremblay, Margaux Simon, Floriane Bretheau, Andréanne K. Blanchette, Catherine Mercier

**Affiliations:** Centre Interdisciplinaire de Recherche en Réadaptation et Intégration Sociale, Quebec City, QC, Canada; Département de réadaptation, Faculté de médecine, Université Laval, Quebec City, QC, Canada; TOPMED, Centre collégial de transfert de technologie en orthèses, prothèses et équipements médicaux, Québec city, QC, Canada

**Keywords:** Locomotion, Task-oriented training, Corticospinal tract, Stroke, Neuroplasticity

## Abstract

**Background:** Corticospinal excitability is subject to alterations after stroke. While the reversal of these alterations has been proposed as an underlying mechanism for improvement walking capacity after gait-specific training, this has not yet been clearly demonstrated. Therefore, the objective of this review is to evaluate the effect of gait-specific training on corticospinal excitability in stroke survivors.

**Design:** Systematic review of the literature

**Methods:** We conducted an electronic database search in four databases (*i.e*., Medline, Embase, CINAHL and Web of Science) in June 2022. Two authors independently screened and selected all studies that investigated the effect of gait-specific training in post-stroke individuals on variables such as motor-evoked potential amplitude, motor threshold, map size, latency, and corticospinal silent period.

**Results:** Nineteen studies investigating the effect of gait-specific training on corticospinal excitability were included. Some studies showed an increased MEP amplitude (7/16 studies), a decreased latency (5/7studies), a decreased motor threshold (4/8 studies), an increased map size (2/3 studies) and a decreased cortical silent period (1/2 study) after gait-specific training. No change has been reported in term of short interval intracortical inhibition after training. Five studies did not report any significant effect after gait-specific training on corticospinal excitability.

**Conclusion:** The results of this systematic review suggest that gait-specific training modalities can drive neuroplastic adaptation among stroke survivors. However, given the methodological disparity of the included studies, further clinical trials with better methodological quality are needed to draw conclusions. Hence, the findings from this review can serve as a rationale for future studies and continued efforts in investigating the effects of gait-specific training on the central nervous system.

## 1. Background

Stroke is a leading cause of physical disability in adults [1]. The prevalence is about 16 million people worldwide [2]. Stroke causes sensorimotor deficits [3,4] that often lead to walking limitations due to the impaired function of neural circuits including the corticospinal tract (CST) [5]. It is recognized that the CST is the main neural pathway that regulates skilled voluntary movement in humans [6,7]. In this context, studies based on non-invasive brain stimulation techniques, such as transcranial magnetic stimulation (TMS) have reported alterations in CST excitability in stroke survivors compared to healthy individuals, such as an increased motor evoked response (MEP) latency [8], an increased resting motor threshold (MT) [9], a reduced MEP size [8,9] and a prolonged silent period [10]. These alterations in CST contribute to alterations motor performance, and are known to be related to gait deficits [11–13]. In fact, compared to healthy individuals, post-stroke individuals often exhibit poor motor control ability [14], reduced walking speed [14,15], frequent falls [16], and limited waking endurance [17]. Because gait limitations prevent their independence in daily activities [18,19], a priority for stroke survivors is to optimize gait recovery [20,21].

Gait-specific training interventions, such as overground training [22], treadmill training without or with bodyweight support [23,24] or robotic-assisted gait training [25,26], focus on the automaticity of walking by providing repetitive stepping practice. These modalities have shown several benefits leading to improve walking ability. Systematic reviews [23,27,28] reported that gait-specific training interventions are beneficial to improve functional / clinical parameters of gait (*e.g*., walking speed, walking endurance, and gross motor function) in individuals with neurological disorders. The functional gains resulting from gait-specific training in stroke survivors, like those produced by gait-specific training, may be due to several mechanisms, such as re-establishing control performed by ipsilesional sensorimotor cortex [29,30] and behavioural compensation strategies [31]. In animals and humans, some studies provided evidence of a change in activation patterns in many regions of the damaged brain [32,33]. Changes in corticospinal excitability might reflect a contribution of primary motor cortex reorganization in functional gains [34,35]. However, although several reviews investigated the effect of interventions on walking capacity in stroke survivors, their impact on corticospinal excitability remains to be clearly established. Therefore, the objective of the present literature review was to summarize and evaluate the effect of gait-specific training on corticospinal excitability in post-stroke individuals.

## 2. Materials and Methods

### 2.1. Data Source and Literature Source

A science librarian was consulted for the initial development of the search protocol. Studies were identified by searching 4 databases (*i.e*., Medline, Embase, CINHAL and Web of Science) from inception to June 2022. The search strategy was based on three main concepts: gait-specific training, corticospinal excitability, and stroke population. More details concerning the search strategy and the keywords used are reported in **Table S1** as supplementary material. The current review follows the guidelines for the preferred reporting items for systematic reviews (PRISMA) [36] and was registered in the PROSPERO register on June 21, 2022 (ID: CRD42022338555).

### 2.2. Eligibility Criteria

The included studies met the following inclusion criteria: (1) intervention studies, such as randomized controlled trials, pre/post studies, case studies, *etc*; (2) studies targeting people who have had a stroke; (3) studies based on gait-specific training modality that focus on practicing specific task related to gait (*e.g*., overground gait training, treadmill training, robotic assisted gait training, *etc*); (4) studies reporting at least one variable related to corticospinal excitability measured with TMS (*i.e*., MEP, motor threshold, map size, latency, and cortical silent period); (5) studies evaluating pre/post effect of the intervention on corticospinal excitability and; (6) studies published in French or English.

Articles were excluded if they: (1) were performed in a mixed population without a possibility to isolate results from individuals who have suffered a stroke; (2) were based on multiple training modalities (*e.g*., including rTMS, transcranial direct-current stimulation, *etc*) among which we cannot distinguish the effects of gait-specific training on corticospinal excitability and (3) were not original research, such as letters to the editor, conference abstracts and commentaries.

### 2.3. Studies of screening

Titles and abstracts of the identified papers were screened independently by two of the authors (YC and AT) to identify those that potentially met the inclusion criteria. A full review of those papers was then performed independently by the same authors. Article selection was discussed until consensus was reached. In the case of any unresolvable disagreement related to the studies eligibility, a third author (CM) intervened to make a decision.

### 2.4. Methodological quality and risk of bias

To assess the methodological quality of included studies, two checklists were used in this study. First, YC and AT independently rated the overall quality of each included article, using the PEDro scale which ranges from 0 to 10 [37]. A PEDro scores of 0-3 were considered ‘poor’, 4-5 ‘fair’, 6-8 ‘good’, and 9-10 ‘excellent’ [38].This checklist allows to identify trials that are likely to be valid and have sufficient statistical information to guide clinical decision-making. Second, the Chipchase checklist was used to evaluate the methodology and reporting of studies in relation to the use of TMS [39]. In this checklist, 8 items are related to subjects (*e.g*., age, gender) and 18 to methodology (*e.g*., coil type, stimulus intensity, *etc*). For both assessment procedures, a calibration meeting was initially held with five articles, to ensure a clear understanding of each criterion and thus standardization and reliability of assessments. A second meeting was held to discuss the criteria for each included article, until a consensus was reached for a score. In the case of any unresolvable disagreement, a third author (CM) performed the assessment to make a decision. For both general PEDro scale and TMS-specific checklist, items were scored as either present (1) or absent (0).

### 2.5. Data Extraction

Data including study design, quality assessment, participants characteristics, intervention (and comparison with a control group), outcomes, and results, was extracted by one author (MS) and validated by a second author (YC). Outcomes of interest were measurements of corticospinal excitability such as MEP size, MEP latency, TMS map area, motor threshold, cortical silent period and short interval intracortical inhibition. In studies in which the gait-specific intervention was a control condition (the experimental condition being for example gait training combined with brain stimulation), the data was extracted only for the pre/post effect of this condition. The quality rating was performed based on a pre/post in such case, therefore reflecting the quality of study based on the data extracted in response to the review objective, and not the quality of the original study.

## 3. Results

### 3.1. Search results

The search and the screening processes are summarized in **Figure 1**. The initial search identified 6174 articles. After removing duplicates, the eligibility of 4008 articles was independently evaluated by two reviewers based on their titles and abstracts. In this process, 82 articles were determined by consensus to qualify for the full-text reading stage. This last stage resulted in the identification of 19 articles as eligible in this review.

**Figure 1.**
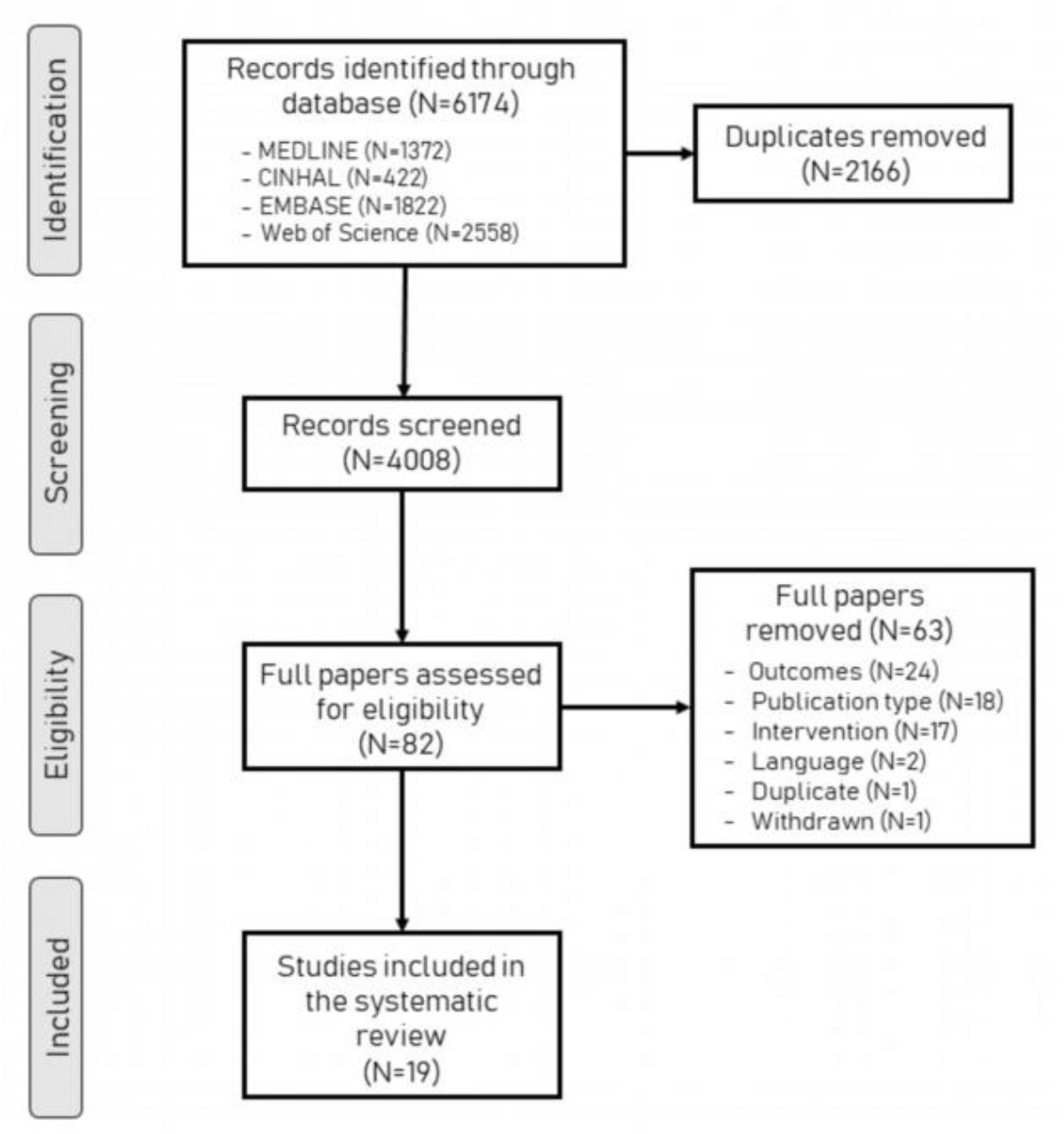
Flowchart outlining the protocol adopted in this systematic review.

### 3.2. Risk of bias

#### Study design and quality assessment

**Table 1** summarize the PEDro rating score for each of the 19 studies, which included 6 randomized controlled trials (RCT) [35,40–44], 2 crossover studies [45,46], 1 cross-sectional study [34], 8 pre/post study [47–53] and 2 cases studies [54,55]. The methodological quality of the included studies ranged from 1 to 8 out of 10, with a median score of 4. Nine studies were of high quality (PEDro score ≥ 6), 6 studies were of moderate quality (PEDro score = 4-5) and 4 were of poor quality (PEDro score ≤ 3). In 6 studies [47–50,52,53], the gait-specific training was considered a control condition to another modality such as tDCS or rTMS. To meet the objective of this review, data extraction in these studies only concerned the pre/post effect of gait-specific training interventions (see Table 1).

**Table 1.** Study design and quality assessment (PEDro score) of the included studies.

| Authors | Design | Item 1 | Item 2 | Item 3 | Item 4 | Item 5 | Item 6 | Item 7 | Item 8 | Item 9 | Item 10 | Item 11 | Total /10 |
| --- | --- | --- | --- | --- | --- | --- | --- | --- | --- | --- | --- | --- | --- |
| Calabrò [40] | RCT | 1 | 1 | 1 | 1 | 0 | 0 | 1 | 1 | 1 | 1 | 1 | 8 |
| Yang [41] | RCT | 1 | 1 | 1 | 1 | 0 | 0 | 1 | 1 | 1 | 1 | 1 | 8 |
| Jayaraman [42] | RCT | 1 | 1 | 0 | 1 | 0 | 0 | 1 | 1 | 1 | 1 | 1 | 7 |
| Li [43] | RCT | 1 | 1 | 0 | 1 | 0 | 0 | 1 | 1 | 1 | 1 | 1 | 7 |
| Shahine [44] | RCT | 1 | 1 | 0 | 1 | 0 | 0 | 1 | 1 | 1 | 1 | 1 | 7 |
| Yen [35] | RCT | 1 | 1 | 1 | 1 | 0 | 0 | 0 | 1 | 1 | 1 | 1 | 7 |
| Forrester [34] | Cross sectional | 1 | 1 | 0 | 1 | 0 | 0 | 0 | 1 | 1 | 1 | 1 | 6 |
| Palmer [45] | Crossover study | 1 | 1 | 0 | 1 | 0 | 0 | 0 | 1 | 1 | 1 | 1 | 6 |
| Li [46] | Crossover study | 1 | 1 | 0 | 1 | 0 | 0 | 0 | 1 | 1 | 0 | 1 | 5 |
| Wang [47]* | Pre/Post study* | 1 | 0 | 1 | 0 | 0 | 0 | 0 | 1 | 1 | 0 | 1 | 4 |
| Chang [49]* | Pre/Post study* | 1 | 0 | 0 | 0 | 0 | 0 | 0 | 1 | 1 | 0 | 1 | 3 |
| Seo [50]* | Pre/Post study* | 1 | 0 | 0 | 0 | 0 | 0 | 0 | 1 | 1 | 0 | 1 | 3 |
| Wang [48]* | Pre/Post study* | 1 | 0 | 0 | 0 | 0 | 0 | 0 | 1 | 1 | 0 | 1 | 3 |
| Wong [51] | Pre/Post study* | 1 | 0 | 0 | 0 | 0 | 0 | 0 | 1 | 1 | 0 | 1 | 3 |
| Koganemaru [52]* | Pre/Post study* | 1 | 0 | 0 | 0 | 0 | 0 | 0 | 1 | 1 | 0 | 1 | 3 |
| Madhavan [53]* | Pre/Post study* | 1 | 0 | 0 | 0 | 0 | 0 | 0 | 1 | 1 | 0 | 1 | 3 |
| Poydasheva [56] | Pre/Post study | 1 | 0 | 0 | 0 | 0 | 0 | 0 | 1 | 1 | 0 | 0 | 2 |
| Krishnan [54] | Case study | 0 | 0 | 0 | 0 | 0 | 0 | 0 | 0 | 1 | 0 | 1 | 2 |
| Peurala [55] | Case study | 0 | 0 | 0 | 0 | 0 | 0 | 0 | 0 | 1 | 0 | 0 | 1 |

#### TMS methodological quality

The specific details of the included studies, which incorporate the evaluation of the Chipchase checklist, are summarized in **Table 2**. The included studies had scores ranging from 5 to 20 out of 26, with a median score of 14. Regarding participant factors, 1 study reported prescribed subject medication (Item 4) and five studies described participants medical condition (Item 7). Concerning methodological factors, the majority of studies reported coil location and stability (N=14), current direction (N=7) or method for determining MEP size during analysis (N=12). Only 3 studies described stimulation pulse shape (N=3) and no studies controlled the level of relaxation present in the muscles other than those being tested.

**Table 2.**
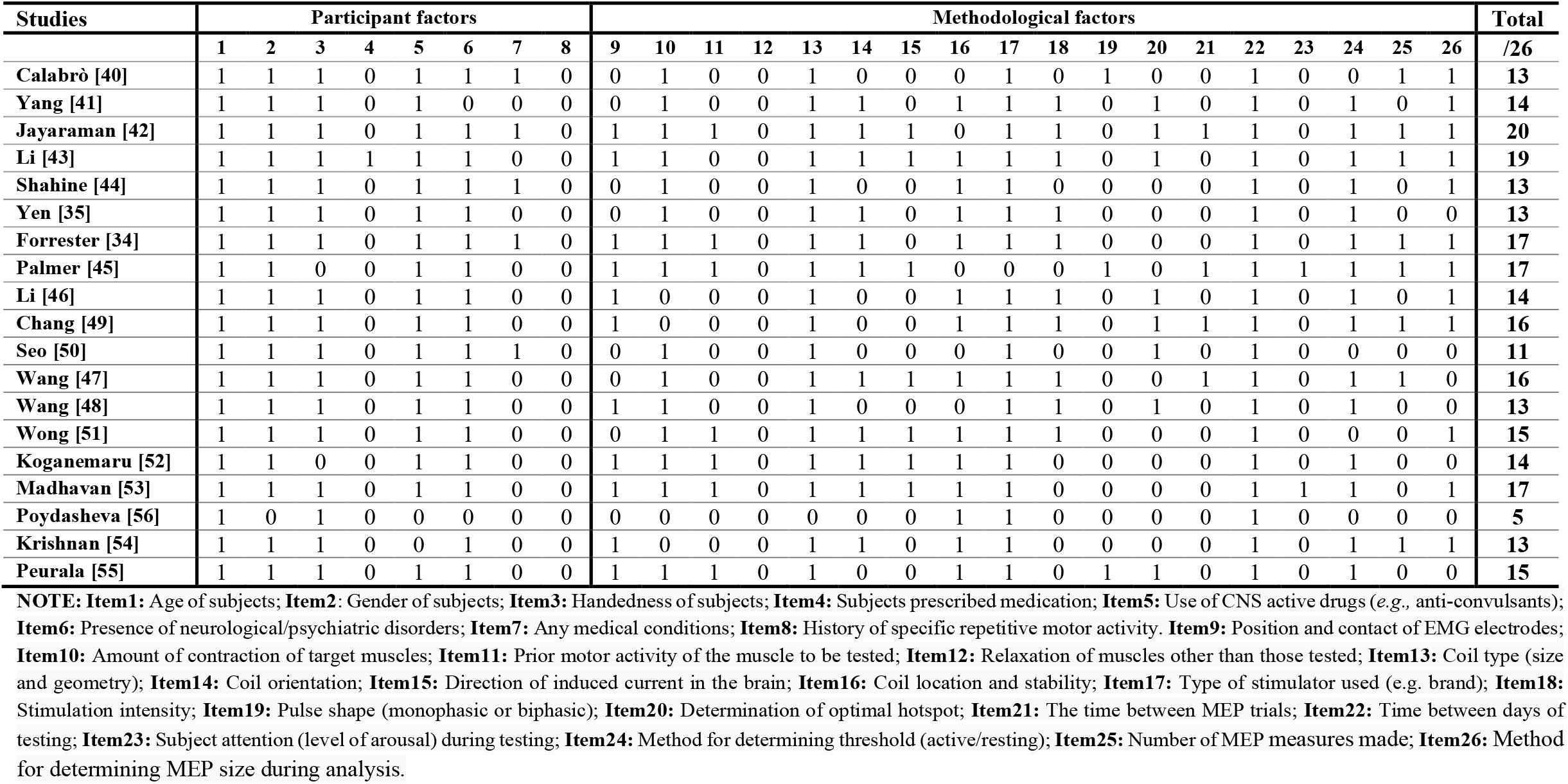
TMS-specific components of methodological quality using the Chipchase checklist.

### 3.3. Characteristics of the participants

The sample size in the included studies ranged from 1 to 50 participants (total of 362 across all studies), and participant demographics varied considerably (see **Table 3**). Sixteen studies focused on participants in the chronic phase of recovery (>3 months post-lesion) after stroke, three studies [41,43,49] included participants in the subacute phase and one study [55] included one participant in the acute phase.

**Table 3:**
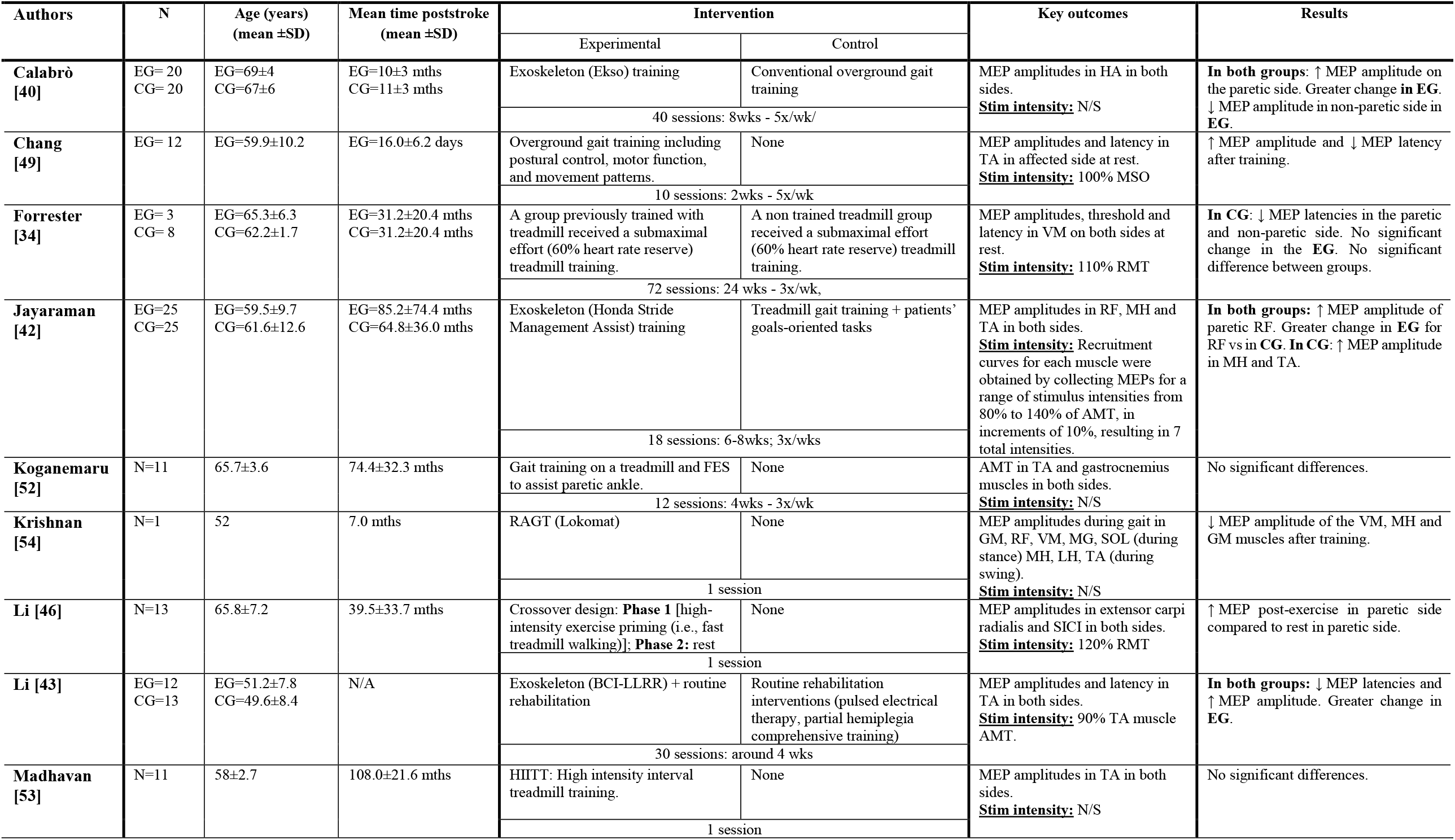
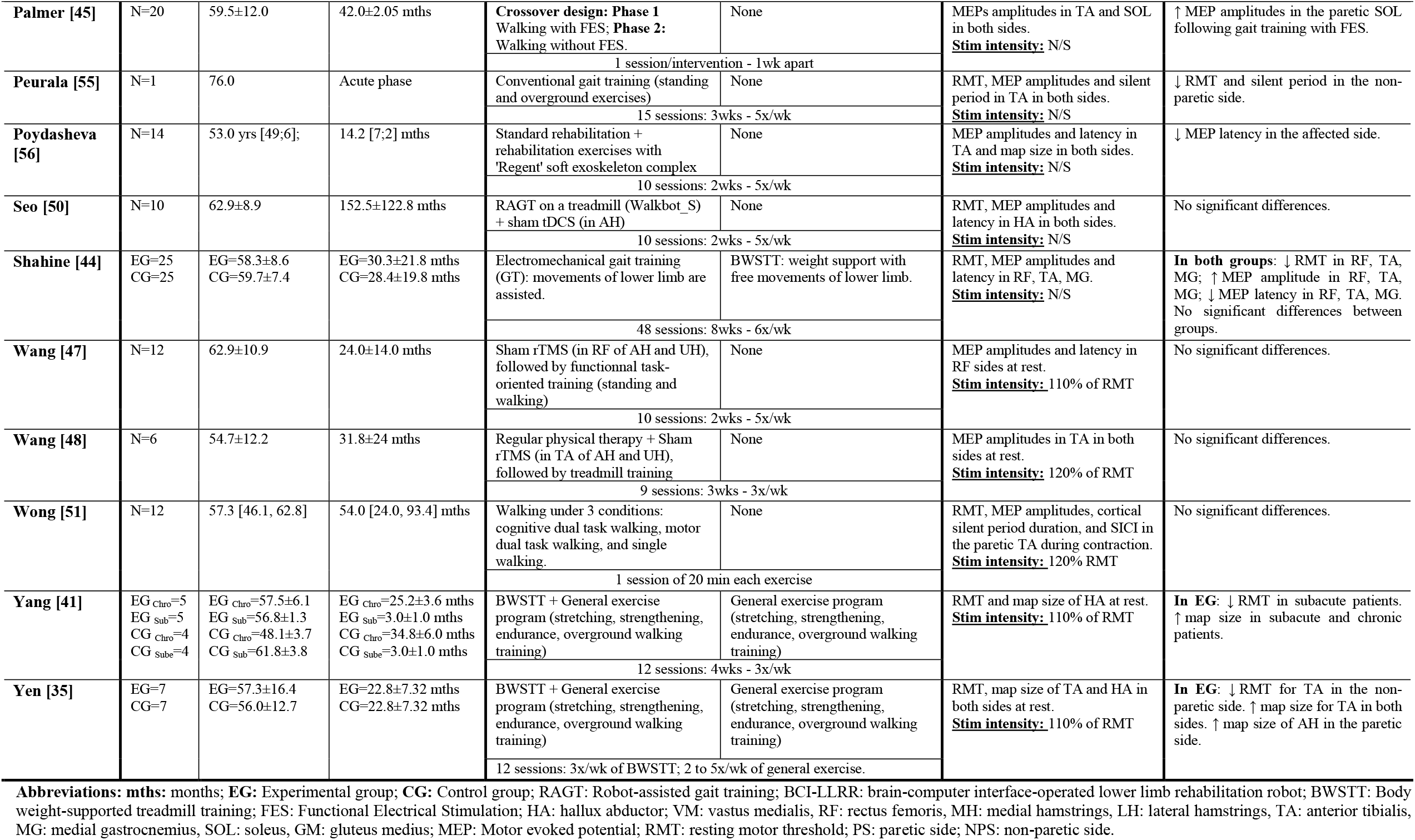
Summary of studies: populations, interventions, outcomes and results.

### 3.4. Gait training protocols

Training parameters (modalities, frequency, session duration and total number of sessions) are displayed in **Table 3**. The protocols of the included studies were heterogeneous (*e.g*., duration: 1 – 24 weeks; frequency: 1 – 5 sessions/week). Of the 5 studies proposing 1-training session, only two studies showed positive effects following gait-specific training (HIIT [46] or walking with FES [45]). However, the majority of studies reporting positive effects were based on protocols with higher training volume (≥12 sessions).

### 3.5. Effect of gait-specific training on corticospinal excitability

The results extracted from the included studies are summarized in **Figure 2** and **Table 3**. The following sections outline the effect of gait-specific training on corticospinal excitability in terms of MEP amplitude and latency, motor threshold, map size, cortical silent period, and SICI.

**Figure 2.**
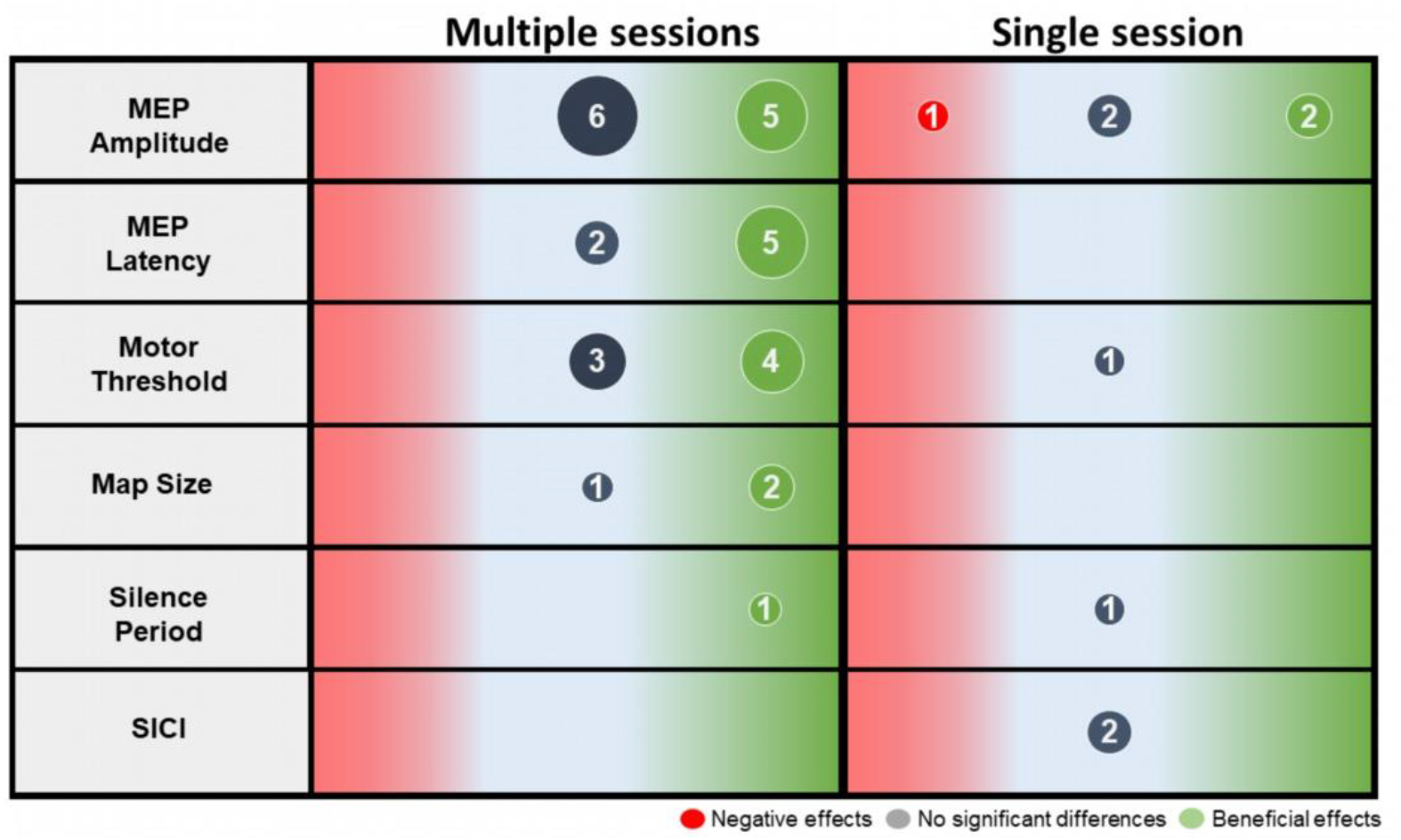
Synthesis of the pre-post effect of single and multiple gait training sessions on corticospinal excitability by reporting the number of articles showing a positive effect (green), no effect (grey) or a negative effect (red).

#### MEP amplitude

Sixteen studies have investigated the effect of gait-specific training on MEP amplitudes. Seven studies showed a significant increase in MEP amplitudes after gait training (*i.e*., robotic training [40,42–44], treadmill training [46], overground training [49] and FES combined to overground training [12]). Five of these studies [40,42–45] presented high methodological quality and two [46,49] were of moderate quality. However, eight studies did not report significant change [34,47,48,50,51,53,55,56] and one study showed a decrease in MEP amplitudes after gait-specific training [54]. Six of these seven studies were of moderate to low methodological quality. Only one study (case study, PEDro=2) reported a negative effect of gait training on MEP amplitude [54]. In general, an increase in the amplitude of MEP may result from gait-specific training; however, in order to better assess corticospinal excitability other TMS parameters should also be considered to compensate for the variability that may be present in the MEP results.

#### MEP latency

Seven studies investigated the effect of gait-specific training on MEP latency. Data from the five studies indicated a decrease in response to conventional gait training [49], robotic training [43,44,49,56], treadmill training [34] and BWSTT [44]. Three of these studies were RCTs (PEDro=6-7) and two were pre/post studies (PEDro=2-4). On the other hand, two pre/post studies (PEDro=3-4) did not observe a significant change in MEP latency after training [47,50].

#### Motor threshold

MT was reported in 7 studies. Four studies reported a decrease in MT after gait-specific training (*i.e*., overground gait training [55], BWSTT [35,41] and robotic gait training [44]), while 3 studies [50–52] did not report any significant changes after training. Three of the four studies showing a positive effect of gait-specific training on MT are RCT (PEDro <7), while those indicating a lack of change are pre/post studies (PEDro=3-4). In general, a significant decrease in MT has been observed after robotic training [44] or BWSTT [35,41,44]. Moreover, Yang et al. [41] observed a decrease in the MT in subacute group but not in the chronic group after BWSTT.

#### Map size

Three studies used the TMS mapping technique to estimate the effect of gait-specific training on the size of the corticomotor representation [35,41,56]. Two RCT studies (PEDro=7-8) reported an increase in map size after BWSTT training [35,41]. Furthermore, in their pre/post study (PEDro=2), Poydasheva et al. [56] did not observe a significant change in this same parameter. Yang et al. [41] found that this increase in map size after gait training was greater in subacute patients compared to chronic patients. Although the results of the two RCTs appear promising in terms of increase in the size of the map, the sample size of these studies remains limited (≤7 participant per group) which limits the conclusions on this variable.

#### Cortical silent period duration

Two studies examined the effect of gait-specific training on the cortical silent period. One study (PEDro=1) showed a decrease in the cortical silent period after overground training in one participant [55] while the other (PEDro=3) did not observe any change in a group of 12 participants [51]. Although the study reporting a lack of effect had a larger sample size, it is important to bear in mind that it looked at the effect of a single training session. Therefore, the available data are too limited to reach a conclusion on this variable.

#### Short interval intracortical inhibition

Two studies [46,51] explored the effect of 1-session gait training on SICI in 12-13 stroke survivors. No significant changes were observed after training.

## Discussion

This review is a first step towards summarize how responses to a single or multiple sessions of gait-specific training can modulate corticospinal excitability in stroke survivors. In general, gait-specific training may enhance corticospinal excitability in stroke survivors. However, given the moderate number of RCT and crossover studies and the overall methodological disparity of included studies (PEDro=1-8), further clinical trials should aim for higher quality designs to better understand the corticospinal responses to gait-specific training.

### Effect of gait-specific training on corticospinal excitability

An effective rehabilitation intervention can modulate the way the brain controls movement [57]. Previous studies showed a remapping of movement representations in M1 in animals after effective rehabilitative training of hand movement after a brain injury [58,59]. Reorganization of corticospinal actions by gait-specific training in individuals with neurologic diseases has been shown in previous studies [58,60]. In this systematic review, we focused on the effect of gait-specific training on corticospinal excitability in stroke survivors on TMS-related outcomes. Most of the included studies (16/19) targeted MEP amplitude, which is an indicator of corticospinal excitation [6]. Among these studies, seven studies showed a significant increase in MEP amplitudes after different modalities of gait-specific training (*i.e*., robotic training [40,42–44], treadmill training [46], overground training [49] and FES combined to overground training [12]). Furthermore, the MEP increases after training have been observed in RF [42,44], TA [43,44,46], SOL [45], MG [44], HA [40] and extensor carpi radialis [46] muscles. However, it is important to mention that eight studies did not report significant change [34,47,48,50,51,53,55,56] and one study [54] showed a decrease in MEP amplitudes after gait-specific training. These studies were mostly of low and moderate methodological quality and performed in participants at the chronic stage. Finally, changes in MEP amplitude have been often investigated as it is relatively easy to quantify. However, other TMS variables might offer a better reliability, such as MT and latency [61].

Lower MTs are associated with increased M1 excitability [62]. Interventions such as motor skill training has been shown to reduce MT in humans [63]. In the present review, several clinical trials reported a reduced MT after gait-specific training (*e.g*., robotic training [44] or BWSTT [35,41,44]) in stroke survivors. An important finding was also the reduction in MEP latency after task-specific gait training [34,43,44,49,56]. This variable appears to be an indicator of lower limb impairment and walking limitations [64]. In two RCTs, the authors reported an increase in map size after BWSTT [35,41]. This allows us to conclude that BWSTT may enlarge the cortical motor representation of TA and HA muscles. Furthermore, a case study [55] showed a decrease in the cortical silent period after 12-session of overground training, while the other [51] did not observe any change after 1-training session in a group of 12 participants. Given the sample size and methodological quality of these studies, it is difficult to draw conclusions on this variable. In conclusion, results derived from several studies on the effect of gait-specific training suggest a positive effect on corticospinal excitability in stroke survivors. However, the lack of consistency in the results, the methodological disparity of included studies (*e.g*., differences assessed muscles, intervention durations, *etc*) and methodological shortcomings in the TMS use should be considered.

It is important to emphasize that some studies included in this review [47,48,50–52,54] did not show a significant increase in corticospinal excitability after gait-specific training in stroke survivors. The methodological shortcomings in the use of TMS could explain the disparity in the results of corticospinal excitability [65]. For example, the hot spot is not always well defined and is sometimes optimized for one muscle while the study evaluates several muscles. The lack of significant post-training change may also be due to the protocols of these studies which are based on a low training volume (*e.g*., 1-12 sessions) [66]. It is possible that an initial increase in corticospinal excitability may still increase with several days and weeks of training but will eventually stagnate and decrease as training progresses without additional challenge [67]. Evidences suggested that the efficacy of post-stroke motor rehabilitation is related to the degree to which the neuromuscular system is challenged by repetitive voluntary movement [68,69]. A single gait-specific training session performed was not sufficient to induce short-term effects on corticospinal excitability parameters [51,53,54] in stroke survivors, except for MEP amplitudes [45,46]. One important methodological factor to consider is that in these studies, measurements were taken right after the training session, while they were typically taken on a different day in studies with multiple sessions. Results of studies using a single session might therefore be impacted by factors such as muscle pre-activation or muscle fatigue. Overall, training parameters (*e.g*., intensity, session duration, frequency) need to be decided in an objective manner [70].

### Clinical recommendations

Despite the methodological disparity of the included studies, some clinical recommendations can be derived from this review. Mostly, studies showed changes in corticospinal excitability after high training volume (≥12 sessions) of gait-specific training in stroke survivors. This observation is consistent with the previous recommendations [66,71] that higher training intensities and durations may promote brain plasticity. However, the heterogeneity of samples and the variability of training modalities, frequencies, durations, and intensity complicates the generation of clear recommendation for optimal gait training parameters that enhance corticospinal excitability after stroke. Furthermore, only one study [41] with a small sample size investigated the effect of gait-specific training on corticospinal excitability in relation to stroke duration, the authors observed a greater increase in corticospinal excitability in subacute group than the chronic group after BWSTT. In the other hand, the studies [47,48,50–53] that did not report significant changes were those targeting individuals in chronic phase. This observation supports the recommendations to start rehabilitation as soon as possible after a stroke [72].

Finally, TMS is a valid tool to evaluate the corticospinal excitability, but it is a technique that presents intra-subject variability and there are few studies on the lower limbs and even less during walking, it is therefore important to be more rigorous in its use, in particular by using TMS-specific components checklist [39]. In our review, we found that the results of the PEDro evaluation did not match the Chipchase checklist. Thus, studies with a high PEDro rating score are not necessarily of good quality from TMS methodology perspective. In conclusion, clinical trials with better methodological quality are needed to better understand the corticospinal responses to gait-specific training.

### Limitations

Some limitations in this review must be acknowledged. First, we reported only TMS outcomes to understand the effect of gait-specific training on corticospinal excitability, while other variables such as EMG-EMG or EEG-EMG coherence might also offer some relevant insight [73]. This choice was made to limit the heterogeneity of the results and allow methodological comparisons across studies. Second, the included studies were diverse regarding the population of patients with stroke, especially regarding the wide variation in time since stroke. Third, another limitation of this review concerns restrictions of publication language and type of publication; therefore, a publication bias might be present.

## Conclusion

This review is a first step towards understanding how M1 and corticospinal pathway respond to a single or multiple sessions of gait-specific training. Overall, the results suggest that multiple gait-specific training modalities can drive neuroplastic adaptation among individuals’ post-stroke even in a chronic phase of recovery. Future studies should aim for higher-quality designs and better TMS methodology so that clear recommendations can emerge and be applied in stroke rehabilitation.

## Data Availability

NONE

## Authors’ contributions

**YC** developed the search strategy and methodology for this review, which have been validated by a science librarian and **CM. YC** and **AT** screened the search hits for eligibility and rated the quality of the included studies. **YC** and **MS** extracted and synthesized the relevant data. **YC** wrote the first draft of the manuscript. **CM, AKB** and **FB** performed a major revision of the manuscript. All authors have read and agreed to the published version of the manuscript.

## Acknowledgments

Y.C. receives a fellowship from Fonds de recherche du Québec—Santé (FRQS). M.S. and A.T. are scholars from Centre interdisciplinaire de recherche en réadaptation et intégration sociale (Cirris). C.M. is Emerita Research Scholar of FRQS and the Laval University Cerebral Palsy Research Chair. The authors thank Martine Gagnon (a science librarian at Laval University) for her guidance and advice during the implementation of the research strategy.

